# A cost-effectiveness analysis of front-line treatment strategies in early stage follicular lymphoma

**DOI:** 10.1101/2021.03.24.21254220

**Authors:** Joshua W.D. Tobin, Anna Crothers, Ti Eric Ma, Peter Mollee, Maher K. Gandhi, Paul Scuffham, Greg Hapgood

## Abstract

Recent data suggests the use of radiotherapy alone (RT) in Early-Stage Follicular Lymphoma is declining. Cost-effectiveness analysis of treatments has not been performed. We constructed a partitioning model (15-year horizon) to compare RT, combined-modality therapy (CMT) and immunochemotherapy with rituximab maintenance (ICT+RM) from a PET-staged cohort from the Australian Lymphoma Alliance. Lifetime direct health care costs, quality-adjusted life-years (QALYs) and incremental cost-effectiveness ratios (ICERs) were calculated. AUD $75,000 was defined as the willingness-to-pay threshold (WTP). The direct healthcare costs were: RT $12,791, CMT $29,391 and ICT+RM $42,644. Compared with RT, CMT demonstrated minimal improvement in QALYs (+0.01) and an ICER well above the WTP threshold ($1,535,488). Compared with RT, ICT+RM demonstrated an improvement in QALYs (+0.41) with an ICER of $73,319. Modelling a 25% cost reduction with a rituximab biosimilar led to further ICER reductions with ICT+RM ($52,476). ICT+RM is cost-effective in early stage FL from the Australian taxpayer perspective.

## Background

Follicular lymphoma (FL) is the most common subtype of indolent non-Hodgkin lymphoma (NHL). Advanced-stage FL has a favourable median overall survival (OS) extending beyond 15 years but is considered incurable with conventional immunochemotherapy (ICT)(1). By contrast, approximately 20% of patients present with early-stage FL and are treated with curative intent (2).

Radiotherapy (RT) has traditionally been considered the standard of care for early stage FL based on studies conducted before the widespread use of rituximab (3-6). These studies demonstrated RT is well-tolerated, provides effective local disease control and may be curative in a subset of patients (3). In an early randomised trial, there was no benefit from adding chlorambucil to RT (7). Therefore, RT alone became the standard of care. Despite this, the majority of patients eventually relapse, typically outside the RT field (5, 6). Importantly, these studies were performed before the widespread adoption of ^18^F-fluorodeoxyglucose positron emission tomography-computed tomography (PET-CT). Therefore, some of the relapsed patients may have had distant sites of PET-avidity at diagnosis that would have correctly assigned them as advanced stage FL.

Recent registry data suggests the use of front-line RT alone is declining as treatment shifts to systemic therapy with the development of rituximab (8-10). Rituximab is an anti-CD20 monoclonal antibody that has prolonged OS in advanced stage FL (11). The use of immunochemotherapy (ICT) with rituximab maintenance (RM) in the front-line setting in early stage FL mirrors the treatment approach in advanced stage FL.

In the modern era, the only randomised evidence to guide treatment is the TROG 99.03 trial that reported significantly improved progression-free survival (PFS) with combined modality therapy (CMT: RT +ICT) compared with RT alone (12). Although this study demonstrates potential to improve PFS, those treated with systemic therapy experienced higher rates of toxicity and there was no improvement in OS. Furthermore, cost-effectiveness was not examined in this study. Consequently, the use of CMT in front-line management remains to be universally adopted.

In addition to clinical efficacy, cost-effectiveness is an important but frequently neglected consideration in facilitating informed public-policy decisions for the treatment of early stage FL(13). Rituximab is one of the highest expenditures to public healthcare systems in most developed economies. In the USA, rituximab has consistently been the second highest expenditure Medicare part B drug since 2015, accounting for between USD $1.5 and $1.7 billion annually (US Centres for Medicare and Medicaid services, 2019). Consequently, the financial burden of systemic therapy is an important consideration in early stage FL, particularly as RT alone, the standard of care for decades, is more affordable (13).

Despite the variability in front-line treatments, an economic analysis of the cost effectiveness of treatment approaches has not been performed in early stage FL. The Australian Lymphoma Alliance (ALA) performed a real-world multicentre comparative assessment of management practices in early stage FL that demonstrated that RT alone and CMT had similar PFS, while ICT+RM demonstrated a significantly improved PFS (14). Here, we performed economic modelling based on this dataset and aimed to estimate the cost-effectiveness of front-line treatment with CMT and ICT+RM compared to RT alone for patients with early stage FL.

## Methods

### Model construction

A four-state partitioned survival model was developed to simulate the total costs and benefits of three front line treatment approaches in early stage FL: RT alone, CMT or ICT+RM (immunochemotherapy and 2 years of rituximab maintenance).

The four disease states were: 1) ‘Failure Free Survival’ (FFS) defined as patients who remain in remission with no requirement for second line treatment; 2) ‘Survival with Failure’ defined as the requirement for retreatment for relapsed or progressive disease; 3) ‘Survival with transformation’ defined as the event of clinical suspicion or biopsy-proven transformation to aggressive lymphoma; 4) ‘Death’ defined as death from any cause (Figure 1). Once a patient fails treatment (relapse, progression or transformation), they remain in that state for the rest of life. Incorporated into the model were the costs of each initial treatment strategy, relapse and transformation costs.

**Figure 1.**
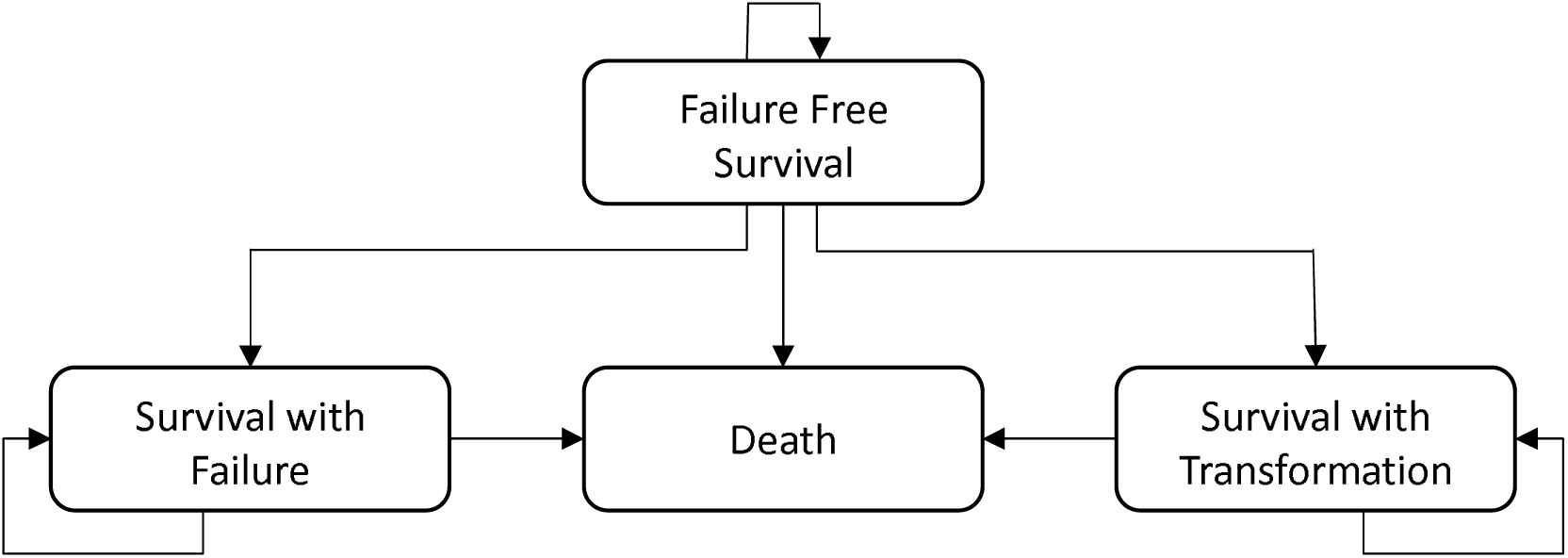
Four-state partitioning model for estimating lifetime health costs of early-stage FL patients.

The probability of moving between health states was evaluated on six-monthly cycles. Simulation was limited to a 15-year time horizon. Treatment arms containing radiotherapy (i.e. RT and CMT) were modelled to have a PFS plateau at 10 years (i.e., no further relapses) representing patients remaining in the failure-free state and presumed to be cured(3, 8, 15). In contrast, the ICT+RM arm was modelled with a stable relapse rate for the entire 15-year simulated time horizon as there are no mature data in the era of PET imaging and rituximab that demonstrates evidence of a cure fraction with non-RT containing approaches. The model assumed: (1) all patients were treated with ICT at relapse; (2) at transformation, all patients were treated with R-CHOP and patients under 65 years also received an autologous stem cell transplantation (ASCT). Outcomes for the baseline analysis were quality-adjusted life years (QALYs) and a cost-utility analysis expressed as an incremental cost-effectiveness ratio (ICER).

### Clinical Data and Transition Probabilities

The clinical data for this economic modelling was derived from an international multicentre retrospective study of patients with early-stage FL from the ALA (14). This cohort included 365 patients with newly diagnosed early stage FL staged with PET-CT and bone marrow biopsy. We excluded patients managed with a ‘watchful waiting’ approach (defined as those not requiring lymphoma therapy for six months from diagnosis), patients receiving ICT only and patients receiving CMT+RM. The economic model was derived from the remaining 242 patients treated actively with RT alone, CMT or ICT+RM (Table 1).

**Table 1.** Clinical characteristics of 242 early stage FL patients from the ALA Cohort.

|  | RT<br>(N = 171) | CMT<br>(N = 34) | ICT + RM<br>(N = 37) | Overall<br>(N= 242) |
| --- | --- | --- | --- | --- |
| Age at Diagnosis (years) | 62 | 54 | 60 | 61 |
| Male | 96 (56%) | 14 (41%) | 23 (62%) | 133 (55%) |
| Female | 75 (44%) | 20 (59%) | 14 (38%) | 109 (45%) |
| Stage I | 125 (73%) | 21 (62%) | 11 (30%) | 157 (65%) |
| Stage II | 46 (27%) | 13 (38%) | 26 (70%) | 85 (35%) |
| Transformation | 11 (6%) | 1 (3%) | 0 (0%) | 12 (5%) |
| Relapse | 42 (25%) | 9 (27%) | 2 (5%) | 53 (22%) |
| Deaths | 12 (7%) | 3 (5%) | 2 (4%) | 17 (7%) |
| Median follow-up (years) | 3.5 | 5.4 | 3.2 | 3.5 |

FFS and OS were estimated by applying Kaplan-Meier estimates for each group until median follow-up. The exponential extrapolation method was uniformly applied for all treatment arms and was selected on the basis of optimal fit for the standard-of-care treatment (i.e., RT) as assessed by the Akaike Information Criterion. In the base case, treatment was assumed to only impact FFS with no impact on OS. This conservative assumption was made to reflect the findings of the TROG 99.03 study which showed no significant OS difference between treatment arms (12).

### Utilities

A utility weight representing health-related quality of life was assigned to each health state. Utility values for each health state were assigned based on a scale of 0 (death) to 1 (perfect health)(Table 2). As utility data was not collected within the ALA cohort, we used published utility values established in patients with advanced stage FL. Patients remaining in the FFS state were assigned to have a utility of 0.88 and patients in the relapsed or transformed disease state were assigned a utility weight of 0.74 (16, 17). Disutilities were applied during cycles in which patients received treatment. No additional disutility was applied during maintenance therapy (18).

**Table 2.**
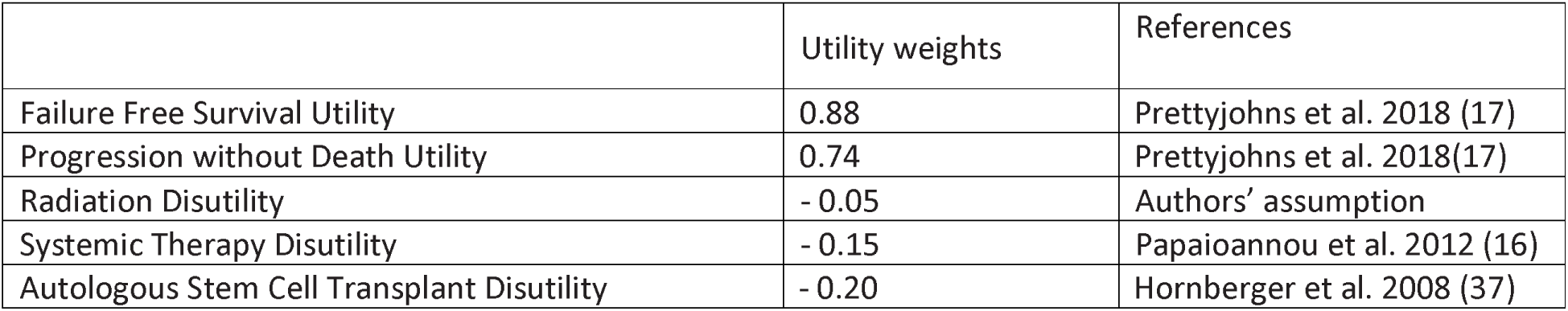
Quality of life adjustments applied in the partitioning model

### Estimating Cost

An Australian tax payer’s perspective was adopted to estimate costs of therapy (Table 3). All costs reflect 2019 values and are reported in Australian Dollars (AUD). The costs of initial work-up and subsequent follow-up were not included as these were likely to be similar among treatment groups. The costs of therapy and associated administration in the outpatient setting were derived from the Australian Pharmaceutical Benefits Scheme and Medicare Schedule (19, 20). Calculations of a chemotherapy dose, and subsequent costs, were based on a patient weight of 70kg with a body surface area of 1.7m^2^. Given the variability of ICT regimens in real world practice (e.g., R-CHOP, R-CVP, R-bendamustine, rituximab monotherapy), we calculated a weighted average cost for treatments. Patients who received maintenance therapy were assumed to receive rituximab every two months over a two-year period (12 cycles)(21). Patients who received RT were assumed to receive 15 fractions (22). The cost of ASCT was derived from the Independent Hospital Pricing Authority (23). Costs associated with hospitalisation from adverse events or palliative care were not included in this evaluation. We performed a series of univariate sensitivity analyses to explore the robustness of the model and the application of clinically relevant assumptions. We altered the value of individual model variables to examine the individual effects on the ICER. In the base-case analysis, annual discount rates of 5% were applied. Cost effectiveness was assessed using a pre-specified willingness-to-pay threshold of AUD $75,000 per QALY gained based on recent approvals in Australia for blinatumomab, brentuximab vedotin and ibrutinib (24-26).

**Table 3.**
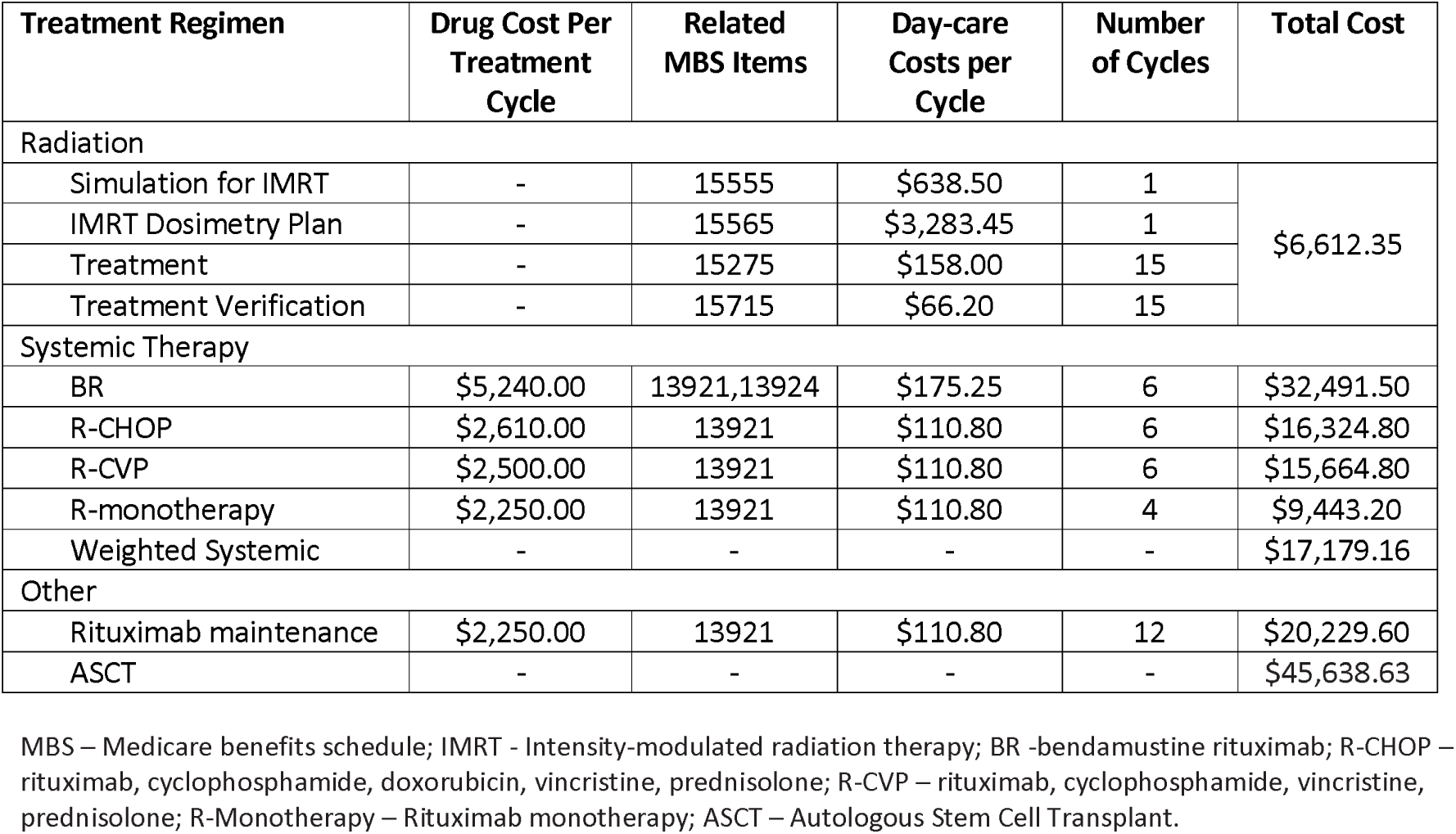
Cost estimates for front-line therapy from an Australian tax payer perspective

## Results

### Patient Characteristics and Clinical Outcomes

The economic model was derived from a total of 242 patients with early-stage FL actively treated with RT alone (n=171), CMT (n=34) or ICT+RM (n=37) in the ALA study. The median follow-up for living patients was 3.5 years (range 0.25-12.8 years). Key patient characteristics are provided in Table 1. The median age was 61 years. There was a similar number of relapses in the RT (n=42, 25%) and CMT groups (n=9, 27%) but significantly fewer events in the ICT+RM group (n=2, 5%)(P=0.009). There was a similar number of transformation events (RT n=11, 6% vs CMT n=1, 3% vs ICT+RM n=0 (P=0.148)) and deaths (RT n=12, 7% vs CMT n=3, 5% vs ICT+RM n=2, 4%)(P=0.880) between all three arms.

### Cost-Effectiveness Analysis

Cost-effectiveness outcomes as predicted by the model with a 15-year time horizon are presented in Table 4. Assuming 5% annual discounting, the total cost of treatment across the time modelled was: RT $$12,791, CMT $29,391 and ICT+RM $42,644. The model demonstrated a shorter time in the failure free health state for both RT (6.51 years) and CMT (6.82 years) compared with ICT+RM (9.41 years). A QALY of 7.85 years was achieved with RT. Compared with RT, no significant difference in QALYs was demonstrated for CMT (7.86; +0.01) yet QALYs were significantly higher for ICT+RM (8.26, +0.41). Compared with RT, the ICER for CMT was $1,535,488 and the ICER ICT+RM was $73,319.

**Table 4.** Comparison of healthcare costs, quality-adjusted life-years and cost-effectiveness between front-line therapies in early stage FL

| Treatment | Cost | | Survival | | QALY | | ICER (\$ per QALY)<sup>1</sup> |
| --- | --- | --- | --- | --- | --- | --- | --- |
|  | Total | Δ Costs<sup>1</sup> | FFS (Yr) | Δ FFS<sup>1</sup> | Total | Δ QALY<sup>1</sup> |  |
| RT | \$12,791 | - | 6.51 | - | 7.85 | - | Reference case |
| CMT | \$29,391 | \$16,660 | 6.82 | +0.30 | 7.86 | +0.01 | \$1,535,488 |
| ICT + RM | \$42,644 | \$29,657 | 9.41 | +2.90 | 8.26 | +0.41 | \$73,319 |

<sup>1</sup> Compared with RT
CMT – combined modality therapy, QALY – quality-adjusted life years; ICER incremental cost-effectiveness ratio (ICER); FFS – failure-free survival; RT – radiotherapy; ICT+RM – immunochemotherapy with rituximab maintenance

### Sensitivity Analysis

Univariate sensitivity analyses were performed on key variables underpinning the model (Table 5). Broadly, all sensitivity analyses resulted in ICER values for CMT patients that remained well above the WTP threshold of $75,000. By contrast, ICT+RM remained relatively stable across all sensitivity variables tested with ICER values remaining below the WTP threshold. The ICT+RM model was sensitive to changes in the time horizon becoming more cost-effective with longer simulated time horizons (15 years = $73,319, 20 years = $62,709). Modelling for a 25% reduction in the cost of RT showed little impact on the ICT+RM ICER value while modelling for a 25% reduction in the cost of rituximab resulted in substantial improvement in the ICER of $52,476 compared to the RT arm.

**Table 5.** Univariate sensitivity analysis of cost-effectiveness model

| | Total costs | | | Total QALY | | | ICER (\$ per QALY)<sup>1</sup> | |
| --- | --- | --- | --- | --- | --- | --- | --- | --- |
|  | RT | CMT | ICT+RM | RT | CMT | ICT+RM | CMT | ICT+RM |
| Base case | \$12,791 | \$29,391 | \$42,644 | 7.85 | 7.86 | 8.26 | \$1,535,488 | \$73,319 |
| Discounting |  |  |  |  |  |  |  |  |
| No Discounting | \$14,063 | \$30,416 | \$44,249 | 10.80 | 10.84 | 11.43 | \$396,494 | \$48,143 |
| Time Horizon |  |  |  |  |  |  |  |  |
| 10 years | \$12,718 | \$29,368 | \$42,644 | 6.24 | 6.23 | 6.52 | Dominant | \$108,815 |
| 20 years | \$12,822 | \$29,401 | \$42,644 | 8.93 | 8.95 | 9.40 | \$625,382 | \$62,709 |
| Cost of radiation |  |  |  |  |  |  |  |  |
| 25% decrease in cost | \$11,137 | \$27,738 | \$42,644 | 7.85 | 7.86 | 8.26 | \$1,535,488 | \$77,379 |
| 25% increase in cost | \$14,444 | \$31,044 | \$42,644 | 7.85 | 7.86 | 8.26 | \$1,535,488 | \$69,260 |
| Cost of rituximab |  |  |  |  |  |  |  |  |
| 25% decrease in cost | \$11,676 | \$25,099 | \$33,043 | 7.85 | 7.86 | 8.26 | \$1,241,586 | \$52,476 |
| 25% increase in cost | \$13,905 | \$33,683 | \$52,246 | 7.85 | 7.86 | 8.26 | \$1,829,390 | \$94,163 |
| Treatment disutility |  |  |  |  |  |  |  |  |
| No disutility | \$12,791 | \$29,391 | \$42,644 | 7.88 | 7.92 | 8.29 | \$373,374 | \$71,454 |
| Treatment for patients who transform |  |  |  |  |  |  |  |  |
| All treated with ASCT | \$12,972 | \$29,449 | \$42,644 | 7.85 | 7.86 | 8.26 | \$1,514,570 | \$72,856 |
| No ASCT | \$12,488 | \$29,295 | \$42,644 | 7.85 | 7.86 | 8.26 | \$1,570,939 | \$74,092 |
| Extrapolation of FFS |  |  |  |  |  |  |  |  |
| Weibull | \$13,887 | \$29,091 | \$42,644 | 7.78 | 7.88 | 8.26 | \$159,957 | \$60,591 |
| Lognormal | \$12,850 | \$28,863 | \$42,644 | 7.84 | 7.89 | 8.26 | \$340,553 | \$71,489 |
| Log-logistic | \$13,210 | \$28,985 | \$42,644 | 7.82 | 7.88 | 8.26 | \$262,401 | \$67,307 |
| Cure model for patients treated with RT and CMT |  |  |  |  |  |  |  |  |
| All groups progress after 10 yrs | \$12,987 | \$29,617 | \$42,644 | 7.82 | 7.83 | 8.26 | \$1,563,918 | \$67,361 |

<sup>1</sup> Compared with RT
CMT – combined modality therapy, ICER incremental cost-effectiveness ratio (ICER); FFS – failure-free survival; RT – radiotherapy; ICT – immunochemotherapy; ICT+RM – immunochemotherapy with rituximab maintenance; ASCT – autologous stem cell transplant

## Discussion

Although RT has traditionally been considered the standard of care for early stage FL, practices are highly variable and include CMT and systemic therapy with rituximab maintenance (9). In this cost-effectiveness analysis, we analysed a real-world cohort of early stage FL patients treated in the era of rituximab and PET-CT. Both CMT and ICT+RM had substantially higher initial costs than RT. Modelling CMT treatment demonstrated only a small increase in the time in the failure-free state; however, there was minimal change to the QALYs compared with RT owing to significant disutility in this cohort due to the use of systemic therapy. Resultantly, CMT was not cost-effective with ICER values well in excess of the WTP threshold. By contrast, despite the higher upfront direct healthcare costs of ICT+RM, the longer period spent in the failure-free state resulted in ICER values remaining below the willingness-to-pay threshold. ICT+RM was a cost-effective treatment approach in early stage FL from the Australian tax payer’s perspective.

RT had been the standard of care in early stage FL based on retrospective data and a randomised trial conducted prior to the modern era of rituximab and PET-CT imaging (3, 4, 6, 7, 27). As most relapses occur outside the RT field (15, 22, 28-30), the concept of adding ICT to RT (CMT) was explored in the TROG 99.03 trial (12). It demonstrated that CMT improved PFS compared with RT but did not show an OS benefit. Furthermore, the addition of ICT was associated with more acute toxicity and the possibility of long-term toxicity. Notably, despite the costs of local versus systemic therapy being so disparate, the cost-effectiveness of treatment strategies has not been previously examined in early stage FL.

Our analysis was based on a retrospective multicentre study where all patients were staged with PET-CT and managed in the rituximab era. This dataset represents the largest study comparing outcomes in PET-staged early stage FL patients. This data showed that, compared to RT alone, CMT had a similar PFS yet patients treated with ICT+RM had an improved PFS. An important caveat is that treatments were not randomly assigned. Unknown confounding variables among patients in treatment groups may have affected results.

In this economic analysis, we examined the cost-effectiveness of CMT and ICT+RM compared to RT. The modelled failure-free survival curves demonstrated no significant difference between RT and CMT, consistent with the non-extrapolated cox-regression analysis in the ALA cohort. As expected, with similar efficacy yet significant disutility with the addition of ICT to RT, CMT was not cost-effective compared to RT. By contrast, although the direct healthcare costs of ICT +RM ($42,644) were higher than RT ($12,791), this was offset by a longer period in the failure-free state over a 15-year simulation. This occurred with modelling for a cure fraction for the RT group and no cure for the ICT+RM group. The high utility in the failure-free state largely explains the ICER values remaining below the willingness-to-pay threshold, despite higher upfront costs. The costs of retreatment for relapse or transformation in these patients were less influential factors in determining the ICER. The base case conclusions of cost-effectiveness regarding both CMT and ICT+RM remained unchanged in the sensitivity analyses. The model was most sensitive to changes in the time horizon. This was due to treatment costs being accrued in the earliest cycles of the model, while the incremental benefit of FFS for ICT+RM was accrued over time.

The costs of treatment approaches in early stage FL are broadly reducing over time. The planning and delivery of RT in indolent lymphoma continues to be refined. Lower doses of RT (24Gy over 12 fractions) have been shown to be non-inferior to higher doses (45Gy)(22). Patients in the ALA cohort were treated with 24-30 Gy with a median of 15 fractions. We examined the impact of a reduction in the cost of RT to reflect this and other efficiencies that may result in the cheaper delivery of RT. The base-case findings were similar with the model being relatively insensitive to changes in the cost of RT.

Similarly, a significant component of the cost of systemic therapy was rituximab (one dose = $2,250). We examined the impact of a reduction in the cost of rituximab as this has fallen significantly since the introduction of biosimilar agents (31). The cost-effectiveness of ICT+RM (ICER $52,467) improved considerably with a 25% reduction in the cost of rituximab in the model. Furthermore, a 25% price reduction is conservative based on the estimates for other biosimilars in Australia (32). The actual price reduction may be greater; however, due to commercial-in-confidence arrangements, this information is not publicly available. This saving is also feasible with no price reduction and the use of 3-monthly maintenance intervals rather than 2-monthly (33). Consequently, the cheaper rituximab becomes, the more cost-effective ICT+RM therapy will be.

The model was designed to reflect an Australian tax payer’s perspective in assessing cost-effectiveness. In Australia, lymphoma treatment is universally available through a publicly funded national health care system. This enabled the determination of the costs of therapy and delivery from national resources (19, 20). Although the costs may differ internationally, the results remain broadly generalisable given the costs incurred related to time in the failure-free state.

Several assumptions were made in our model. Consistent with current practice, we assumed that at transformation, patients <65 years would proceed to ASCT after ICT, while patients >65 years would receive ICT only (34). Due to the similar rates of the rare event of transformation between groups, the model remained largely unchanged by changes to this assumption on univariate sensitivity analysis. Costs associated with adverse events, which are more common in systemic treatment, were not included. RT is well tolerated with low rates of early and late toxicity (12, 29). Conversely, systemic therapy is associated with more frequent acute toxicity (12); however inpatient or supportive care of adverse events is uncommon and economic modelling of similarly-treated patients with advanced-stage FL has shown these adverse events contribute minimally to direct healthcare costs of therapy (35). The impact on cost-effectiveness of both short and long-term toxicities is often burdened by decreased quality of life, which is reflected in the disutility assigned to both the ‘failure’ health state and the disutility applied in the simulation during treatment periods. We used an average cost of commonly used systemic regimens (BR, R-CHOP, R-CVP and rituximab monotherapy) reflecting variable clinical practice and similar PFS among therapies from previous datasets (9, 14).

To our knowledge, this is the first economic analysis of front-line treatment approaches in early stage FL. Although the TROG 99.03 study demonstrated a PFS advantage with CMT compared to RT alone, other measures, such as cost and toxicity, are pertinent when considering the widespread application of this approach. Our data demonstrated that the significant initial cost of ICT+RM was offset by improvements in FFS and subsequently improved utility over the long-time horizon experienced in these patients. This suggests that ICT+RM can be cost-effective despite the substantially higher upfront costs. Rituximab monotherapy is a promising candidate in conjunction with RT and currently under investigation in a randomised trial in early stage FL(36). Modelling a conservative 25% price reduction for rituximab with the introduction of biosimilars, ICT+RM was more cost-effective, demonstrating that the cheaper rituximab becomes, the more cost-effective systemic therapy will be. Future clinical trials in early stage FL should include measures of cost-effectiveness to facilitate public-policy and reimbursement decisions.

## Data Availability

Data available on request

## Authorship

### Conception and design

JWDT, MG, GH.

### Collection and assembly of data

JWDT, AC, EM, MG, PS, GH

### Data analysis and interpretation

JWDT, AC, MG, PS, GH

### Manuscript writing

All authors

### Final approval of manuscript

All authors

